# Real-life Psychological Experiences and Subsequent Cardiovascular Events among Survivors of a Myocardial Infarction in Midlife: Prospective Cohort Study

**DOI:** 10.64898/2026.09.30.26364457

**Authors:** Viola Vaccarino, Hua She, Lisa Elon, Tené T. Lewis, Yi-An Ko, Nancy Murrah, Lucy H. Shallenberger, Tatum Roberts, Lewam Stefanos, Jose Medina Inojosa, Kristen Harris, Taha Ahmed, Alexander Razavi, J. Douglas Bremner, Paolo Raggi, Arshed Quyyumi, Amit J. Shah

## Abstract

**Background:** Information is lacking about everyday psychological states of individuals with a recent myocardial infarction (MI) and whether they influence outcomes. We investigated real world psychological experiences and their association with subsequent cardiovascular events in survivors of an MI.

**Methods:** We studied 287 individuals ≤ 60 years of age within 8 months from an MI. At the baseline visit, we assessed sociodemographic and clinical factors, and administered a battery of psychological scales of depression, anxiety, anger, perceived stress and social support. Participants underwent a 7-day home monitoring of mood and stress using ecological momentary assessments, and a momentary negative affect (MNA) score, integrating daily mean ratings of sadness, anxiety, anger, distress and loneliness, was calculated. Participants were followed for a median time of 28 months. The study endpoint was a composite index of major adverse cardiovascular events (MACE), including cardiovascular death, nonfatal MI, stroke or hospitalizations for heart failure. Cox proportional hazards models adjusted for sociodemographic and clinical factors.

**Results:** The mean age of the sample was 51 years, and 43% of participants were women. Each standard deviation increase in MNA was significantly associated with a 60% increased risk of MACE (95% confidence interval (CI), 20% to 100%), irrespective of adjustment for sociodemographic factors and clinical characteristics. The association was similar in women and men. Momentary positive mood and momentary perceived stress were not associated with MACE. The MNA score showed a larger independent contribution to MACE compared to the combined battery of baseline psychological assessments and the association was independent of baseline history of depression.

**Conclusion:** Among individuals after MI, moment-to-moment negative affect during one week of monitoring in daily life is a powerful predictor of subsequent cardiovascular events. A prospective evaluation of negative mood during everyday living could represent a novel strategy to uncover residual risk after MI.

---

Negative affect, encompassing mood states such as depression, anxiety, anger, perceived stress, and loneliness, is well documented to influence the onset, recovery, and outcome of cardiovascular disease (CVD).^1,2^ This relationship is especially evident among people with pre-existing CVD,^3,4^ among whom negative affect can also impair post-discharge medication adherence and rehabilitation.^5,6^ These observations, however, are mostly based on retrospective self-reports collected at a single point in time. Alternatively, some studies have used laboratory models of stress provocation.^7^ Both approaches have uncertain accuracy for capturing the true frequency of such exposures in everyday life, which limits their relevance to disease pathophysiology and outcome.

Ecological momentary assessment (EMA) offers an alternative to these approaches, capturing repeated, real-time reports of experiences as they occur in participants’ natural environments.^7–9^ Collecting data in real time and in the real-world enhances ecological validity and avoids the pitfalls of retrospective report. EMA may also help identify which psychological experiences confer risk and clarify how these effects unfold over the course of daily life. EMA protocols have shown high adherence and feasibility in studies of predominantly healthy samples^10^ as well as in patients with acute coronary syndromes,^11^ including our experience with individuals after an MI.^12^

EMA of negative and positive mood, along with other dimensions of psychological distress, has been used previously in community samples in relation to CVD risk factors and subclinical CVD.^7,13^ However, data remain scarce in high-risk groups such as individuals after an MI. Recently, using EMA we showed that women post-MI have more negative affect during everyday life than men, which was related to worse ambulatory autonomic physiology.^12^ However, whether negative affect in daily life is related to adverse outcomes in survivors of an MI is unknown. The purpose of this study was to assess whether real-life negative affect, assessed with EMA three times daily over one week of monitoring, is related to subsequent CVD outcomes in people who recently survived MI. We further examined whether real-life assessments are more informative in predicting outcomes than psychological scales administered at the baseline visit. We also contrasted daily-life negative affect with daily life positive affect assessed in a similar fashion.

## Methods

### Study sample

This report is based on participants enrolled in the Myocardial Infarction and Mental Stress Study 3 (MIMS-3), recruited from Emory University-affiliated hospitals across metropolitan Atlanta between February 2020 and December 2023.^12^ Eligible individuals were between 18 and 60 years of age at the time of their MI and had a documented hospitalization for type 1 MI within the preceding 8 months.^14^ Type 1 MI diagnosis was confirmed through medical record review using published criteria of elevated troponin levels, which had to be accompanied by ischemic symptoms and/or new ischemic electrocardiographic changes or development of pathological Q waves or other evidence of myocardial necrosis or atherothrombosis.^14^ We excluded individuals with a serious comorbid medical or psychiatric condition that might interfere with participation or follow-up, including cancer, renal failure, poorly controlled hypertension, active alcohol or substance abuse, or schizophrenia, as well as those who were pregnant or breastfeeding or taking immunosuppressant or psychotropic medications other than antidepressants. All procedures were approved by the Emory University Institutional Review Board and every participant provided written informed consent.

### Baseline Assessments

At the baseline clinic visit, trained staff recorded sociodemographic characteristics, medical history, and current medication use, and measured height and weight to calculate BMI. Race was self-identified. A battery of validated instruments assessed behavioral, social, and health-related domains. Lifetime major depressive disorder was ascertained using the Structured Clinical Interview for DSM-IV (SCID),^15^ administered by research staff who were trained and directly supervised by the study psychiatrist (JDB). Current depressive symptoms, referring to the prior 2 weeks, were measured with the Beck Depression Inventory-II.^16^ Additional baseline measures included the Spielberger’s Anxiety Inventory (20-item state anxiety module),^17^ the Spielberger’s Anger Inventory (15-item state anger module),^18^ the Perceived stress Scale, 4-item version,^19^ and the Multidimensional Scale of Perceived Social Support, a 12-item self-report questionnaire designed to measure a person’s subjective perception of social support from three specific sources: family, friends, and a significant other.^20^ To derive a single baseline index of negative affect for comparison against momentary experiences reported in daily life, we standardized (mean 0, standard deviation (SD) 1) and summed the scores across these five instruments. Because a higher score for the Multidimensional Scale of Perceived Social Support indicates better social support, the score was reversed before standardization.

Clinical and angiographic details of the index MI were abstracted from medical records by trained staff. Obstructive CAD was defined as luminal stenosis of 70% or greater. CAD severity was summarized with the Gensini score, which weights the degree of stenosis by the prognostic importance of its location within the coronary tree.^21^

### Monitoring of Daily Psychological Experiences

Participants used a study-provided smartphone, or their own device if preferred, to complete ecological momentary assessments (EMA) of everyday perceived stressors and mood states, adapting methodology from the National Study of Daily Experiences component of the Midlife in the United States Study.^22^ Surveys were delivered through a smartphone application (RealLife Exp; LifeData LLC, Marion, IN) that prompted participants three times a day for 7 consecutive days at semi-random intervals—once between 10 AM and 1 PM and twice between 2 PM and 10 PM—each separated by a minimum of 180 minutes. At each prompt, participants rated, on a 0-to-100 scale (none of the time to all of the time), and with reference to the preceding hour, their levels of sadness, anxiety, anger, distress, loneliness, happiness and calmness, and indicated, with a yes/no response, whether anything had occurred in that hour that most people would find stressful.^23^

The primary exposure, a momentary negative affect (MNA) score ranging from 0 to 100, was derived by averaging the five negative-emotion ratings (sadness, anxiety, anger, distress, and loneliness) within each day and then averaging those daily values across the week; this two-step approach allowed participants with incomplete monitoring days to be retained. An analogous momentary positive affect (MPA) score was calculated from the weekly average of daily happiness and calmness ratings. Within-person mood fluctuation was captured as the average number of standard deviations of negative and positive ratings over the week, and momentary stress was defined as the proportion of prompts across the week at which a perceived stressor was reported.

### Outcome Assessment

After the baseline visit, participants were prospectively followed through approximately annual contacts, with follow-up extending through May 31, 2026. At each contact, study staff ascertained cardiovascular events and hospitalizations through direct participant contact, medical record abstraction, and searches of vital records; reported hospitalizations or procedures prompted outreach to the treating physician and retrieval of hospital records. All events were adjudicated by study cardiologists unaware of participants’ other study data. The primary outcome was a composite of major adverse cardiovascular events (MACE), including cardiovascular death, nonfatal MI, stroke, or hospitalization for heart failure.

### Statistical analysis

Our main predictor was the MNA score, the daily average of the 5 ratings of negative affect across the week. We first assessed whether the association between the MNA score and MACE departed from linearity, using restricted cubic splines with knots at the 25th, 50th, and 75th percentiles (**Supplemental Figure 1**). Nonlinearity in the association between the MNA score and MACE was assessed using a likelihood ratio test comparing the restricted cubic spline model to a model in which the MNA was entered as a single linear term. Since this analysis did not indicate significant departure from linearity (LR χ²(1) = 0.26, p = 0.61), the MNA score was primarily analyzed as a continuous variable after standardization to a mean of 0 and a SD of 1. For descriptive purposes, demographic, behavioral and clinical characteristics were tabulated according to the MNA score categorized at its median value; no hypothesis testing was done in this descriptive analysis.^24^

To examine the relationship between daily life psychological experiences and CVD events we used Cox proportional hazards models. The primary dependent variable was the MNA score as a continuous variable, scaled as SD increase. We adjusted for baseline variables deemed *a priori* the most important potential confounding factors, including demographic factors (age, sex, race, education, marital status), and clinical risk factors and severity indicators (ever smoking, body mass index [BMI], history of hypertension, history of diabetes mellitus, left ventricular ejection fraction, and ST-segment elevation MI [STEMI], vs. non-STEMI, as the index event). Similar analyses were conducted for the MPA score, for mood fluctuation, and for daily perceived stress. In all the models we tested the interaction between daily psychological experiences and sex. To improve clinical interpretation, we also repeated the multivariable analysis with the MNA score dichotomized at the median value.

Because the number of outcome events was small relative to the number of covariates, we performed a sensitivity analysis using propensity score methods to reduce the dimensionality of confounder adjustment and minimize the risk of model overfitting.^25^ For the continuous exposure (the standardized MNA score), we estimated a generalized propensity score as the predicted value from a linear regression model of the exposure on the covariates.^26^ For the dichotomous MNA, the propensity score was estimated as the predicted probability of being above the median from a logistic regression model. Both propensity score models included sex, age, race (Black vs. other), education (>12 years), marital status, ever smoking, body mass index, history of diabetes, history of hypertension, left ventricular ejection fraction, and STEMI. We also constructed separate propensity scores that also included medications (beta blockers, angiotensin converting enzyme inhibitors, angiotensin receptor blocker medications, statins and antidepressants). Models included the exposure and the propensity score, the latter modeled flexibly as a restricted cubic spline with 4 knots placed at the 20th, 40th, 60th, and 80th percentiles.

To quantify the relative importance of moment-to-moment psychological states vs. assessments conducted retrospectively at one point at the baseline visit, we used several approaches. First, we examined the association of individual components of the MNA score, i.e., daily mean ratings of sadness, anxiety, anger, distress and loneliness, with MACE, and contrasted their association with that of negative affect scales assessing similar dimensions at baseline, including depressive symptoms, state anxiety, state anger, perceived stress, and social support, as well as their combination in the baseline negative affect score which summed the scores across these five instruments after standardization. In all these analyses, the variables of interest were standardized (mean 0, SD 1) prior to model fitting so that their scales would be comparable.

Second, we constructed joint models that adjusted for the MNA score and the baseline negative affect score in the same model. Third, we characterized the relative importance of these two scores in predicting outcomes. Specifically, we calculated the partial likelihood ratio chi-square statistic for each variable from the multivariable Cox proportional hazards model, comparing the −2 log partial likelihood of the full model to that of a reduced model omitting only that covariate. The resulting difference, distributed as a chi-square statistic with 1 degree of freedom, reflects the variation in the likelihood of the outcome uniquely attributable to that covariate after adjustment for the remaining covariates in the model. This statistic also allowed us to rank variables in the model by importance, reflecting each variable’s statistical contribution to model fit. In a sensitivity analysis we substituted the global baseline negative affect score with the baseline assessments of depression, including the Beck Depression Inventory score and lifetime history of major depression.

The proportional hazards assumption was evaluated using martingale residual-based diagnostics implemented via the ASSESS statement in PROC PHREG. This analysis showed that there was no deviation from the proportionality assumption. Missing covariate data were minimal (<4%). Therefore, we did not perform multiple imputations. All analyses were conducted using SAS statistical software version 9.4 (SAS Inc., Cary, NC).

## Results

### Study sample

Of the 306 participants enrolled in the MIMS-3 study, 287 had EMA data and were included in the analysis. The sample included 123 women and 164 men; the mean age was 51.3 years (SD, 7.1 years), with an age range of 26 to 60 years. The weekly average percentage of EMA prompts that were filled over the maximum expected was 75.1% (SD, 25.7%). The median time from the date of the index MI to the study visit was 3.0 months (interquartile range, 2.5 months).

Individual components of the MNA, including daily mean ratings of sadness, anxiety, anger, distress and loneliness, were significantly correlated with each other and with reports of stress during the week (**Supplemental Table 1**). The mean MNA score, on a scale of 0 to 100 before standardization, was 12.1 (SD, 14.8), with a median of 6.1. The mean MPA score, integrating daily mean ratings of happiness and calmness, on a scale of 0 to 100, was 73.1 (SD, 21.6), and the mean momentary mood fluctuation across the week was 5.6 standard deviation units (SD, 4.5). The mean percent of moments with stress during the week was 13.8% (SD, 16.2%). The percent of EMA prompts that were completed was weakly negatively correlated with the MNA score (Spearman r=-0.22, p<0.001) but not with the MPA score (Spearman r=0.07, p=0.25) or the percent of moments with stress during the week (Spearman r=-0.04, p=0.44).

### Baseline Characteristics

Participants with a MNA score greater than the median value of 6.1 tended to be younger, more often female, less often married and with lower income despite similar levels of education (**Table 1**). However, CVD history and clinical characteristics were similar, as was medication use, except for antidepressant use that was more common in participants with higher MNA score. Participants with higher MNA score also had a higher lifetime prevalence of major depression at baseline, and higher scores of baseline psychological scales (depression, anxiety, anger, and perceived stress), while scale scores for social support were lower.

**Table 1.** Descriptive characteristics of the study population by the momentary negative affect score categorized at the median value.

|  | MNA ≤Median<br>(0 to 6.1)<br>N=143 | MNA > Median<br>(6.2-76.1)<br>N=144 | N Missing |
| --- | --- | --- | --- |
| **Demographic Factors** |  |  |  |
| Age, years, mean (SD) | 53 (6) | 50 (7) | 0 |
| Female, % | 38 | 48 | 0 |
| Black or African American, % | 50 | 56 | 0 |
| Married or living with partner, % | 54 | 48 | 0 |
| Education >12 years | 69 | 67 | 0 |
| Income < \$25,000, % | 17 | 29 | 1 |
| **Cardiovascular History and Risk Factors** |  |  |  |
| BMI, kg/m<sup>2</sup>, mean (SD) | 31 (6) | 31 (6) | 0 |
| Ever smoker, % | 39 | 42 | 1 |
| Systolic blood pressure, mmHg, mean (SD) | 132 (18) | 132 (21) | 0 |
| Diastolic blood pressure, mmHg, mean (SD) | 82 (11) | 82 (13) | 0 |
| Heart rate, beat/min, mean (SD) | 63 (11) | 65 (13) | 0 |
| History of hypertension, % | 75 | 79 | 0 |
| History of dyslipidemia, % | 72 | 76 | 0 |
| History of diabetes, % | 34 | 36 | 0 |
| History of MI prior to index event, % | 16 | 23 | 5 |
| History of revascularization, % | 87 | 93 | 0 |
| **Clinical Characteristics** |  |  |  |
| LV ejection fraction (%), mean (SD) | 49 (12) | 50 (12) | 0 |
| ST-segment elevation MI, % | 30 | 28 | 0 |
| Gensini CAD severity score, mean (SD)* | 32 (4) | 29 (4) | 4 |
| Obstructive CAD (stenosis ≥70%), % | 85 | 83 | 4 |
| Three-vessel disease (at ≥70%), % | 9 | 11 | 4 |
| **Medications** |  |  |  |
| Beta Blocker, % | 85 | 87 | 1 |
| Statins, % | 89 | 88 | 0 |
| Aspirin, % | 87 | 89 | 1 |
| ACE Inhibitors, % | 32 | 24 | 1 |
| ARB medications, % | 32 | 24 | 0 |
| Antidepressants, % | 9 | 20 | 1 |
| **Baseline Psychological Factors** |  |  |  |
| Lifetime history of major depression, % | 18 | 50 | 3 |
| Beck Depression Inventory, mean (SD) | 7 (6) | 14 (10) | 0 |

**Table 1 (Continued)**
|  |  |  |  |
| --- | --- | --- | --- |
| State Anxiety, mean (SD) | 28 (8) | 40 (13) | 8 |
| State Anger, mean (SD) | 16 (2) | 21 (10) | 8 |
| Perceived Stress Scale, mean (SD) | 4 (3) | 7 (4) | 3 |
| Multidimensional Scale of Perceived Social Support, mean (SD) | 66 (17) | 58 (18) | 13 |

SD: standard deviation; BMI: Body mass index; ACE: angiotensin-converting enzyme; ARB: angiotensin receptor blocker; MI: myocardial infarction; LV: left ventricle; CAD: coronary artery disease.
\* Geometric means.

### Relationship of Moment-to-Moment Assessments with Cardiovascular Outcomes

After a median follow-up time of 28.0 months (interquartile range, 13.5 months), there were 43 MACE events (22 in men and 21 in women), including 16 nonfatal MIs, 17 heart failure hospitalizations, 5 strokes, and 5 CVD deaths. There was a significant association between the MNA score and MACE, which was robust to the adjustment for demographic factors, CVD history and clinical characteristics (**Table 2**). Irrespective of adjustment (Models 1 to 3), each SD increase in MNA, corresponding to a 15-point increment in the raw scale from 0 to 100, was associated with 60% higher risk of MACE (95% confidence interval (CI), 20% to 100%).

**Table 2.** Association of the momentary negative affect score (MNA) with major adverse cardiovascular events (cardiovascular death, nonfatal MI, stroke or hospitalizations for heart failure).

|  | N in<br>model | Per SD Increment of<br>MNA Score |  | MNA Score ≤ Median vs.<br>MNA Score > Median |  |
| --- | --- | --- | --- | --- | --- |
|  |  | HR (95% CI) | P Value | HR (95% CI) | P Value |
| Main Models |  |  |  |  |  |
| Model 1. Unadjusted | 287 | 1.6 (1.2, 2.0) | <0.001 | 2.5 (1.3, 4.9) | 0.006 |
| Model 2. Adjusted for sociodemographic factors* | 287 | 1.6 (1.2, 2.0) | <0.001 | 2.6 (1.3, 5.1) | 0.005 |
| Model 3. Adjusted for the above plus cardiovascular history, cardiovascular risk factors and clinical characteristics† | 286 | 1.6 (1.2, 2.0) | <0.001 | 3.5 (1.7, 7.4) | <0.001 |
| Sensitivity Models |  |  |  |  |  |
| Model 4. Using a propensity score for covariates in Model 3 | 286 | 1.5 (1.2, 1.9) | 0.001 | 2.9 (1.5, 5.8) | 0.002 |
| Model 5. Using a propensity score for covariates in Model 3 and cardiovascular‡ and antidepressant medications | 284 | 1.5 (1.2, 1.9) | 0.002 | 2.9 (1.4, 5.9) | 0.003 |

MNA: Momentary Negative Affect. HR: Hazards ratio. CI: confidence interval.
One SD corresponds to 15 points of the momentary negative affect score.
The interaction between MNA and sex was not significant in any of the models.
\*Age, sex, race (black versus non-black), married vs. not, education >12 years.
† Ever smoking, BMI, history of hypertension, history of diabetes mellitus, left ventricular ejection fraction, and ST-segment elevation MI.
‡ Beta-blockers, angiotensin-converting enzyme inhibitors, angiotensin receptor blockers, and statins.

Propensity score analysis provided similar results, therefore suggesting that model overfitting was not a concern (Model 4), with estimates that remained similar even after medications were added to the propensity score (Model 5). When we repeated the analysis with the dichotomized MNA score, a MNA score above the median was associated with a 2.5 to 3 times higher risk of MACE (**Table 2**). The association was similar in women and men and the interaction between sex with MNA was not significant in any of the models. After adjusting for demographic and clinical risk factors (Model 3), the hazards ratio (HR) per SD increase in MNA was 1.7 (95% CI, 1.2, 2.5) in women, and 1.5 (95% CI, 1.1, 2.0) in men, p=0.47 for the interaction.

In contrast to the MNA score, the MPA score was not significantly associated with MACE. After adjusting for demographic and clinical risk factors (Model 3), the HR per SD increase in MPA was 0.8 (95% CI, 0.5, 1.1). Moment-to-moment mood fluctuations were also not associated with outcomes (HR, 1.1 per SD increase, 95% CI, 0.7, 1.5). Perceived moment-to-moment stress did not show an association with MACE either: HR, 1.01 (95% CI, 0.99, 1.03) per percentage point increase in moments with stress during the week. These associations also did not vary significantly between women and men.

### Relative Importance of Moment-to-Moment Assessments vs. Baseline Psychological Scales

Baseline psychological scales were correlated with each other (**Supplemental Table 2**), as well as with momentary assessments (**Supplementary Table 3**). Spearman correlation coefficients ranged between 0.2 and 0.6. When examined in relation to outcome, all the individual components of the MNA were significantly associated with MACE after adjusting for demographic factors, medical history and clinical variables (**Table 3**). In contrast, of the baseline psychological scales, only depressive symptoms were significantly associated with outcome (**Table 3**). The global baseline negative affect score, which combined the ratings of baseline scales, was not significantly associated with MACE (HR per SD increase, 1.3, 95% CI, 0.9, 1.9).

**Table 3.** Association of individual momentary ratings of negative affect and of corresponding baseline psychological rating scales with major adverse cardiovascular events (cardiovascular death, nonfatal MI, stroke or hospitalizations for heart failure).

|  | N in<br>model | HR (95% CI) Per SD Increment | P Value |
| --- | --- | --- | --- |
| **Momentary Assessments\*** |  |  |  |
| Sad | 286 | 1.4 (1.1, 1.8) | 0.01 |
| Anxious | 286 | 1.6 (1.2, 2.1) | <0.001 |
| Angry | 286 | 1.5 (1.1, 1.9) | 0.003 |
| Distressed | 286 | 1.4 (1.0, 1.8) | 0.03 |
| Lonely | 286 | 1.3 (1.1, 1.7) | 0.02 |
| **Baseline Assessments** |  |  |  |
| Beck Depression Inventory | 286 | 1.7 (1.2, 2.3) | <0.001 |
| State Anxiety | 278 | 1.3 (0.9, 1.9) | 0.15 |
| State Anger | 278 | 1.0 (0.7, 1.3) | 0.99 |
| Perceived Stress Scale | 283 | 1.2 (0.8, 1.8) | 0.46 |
| Multidimensional Scale of Perceived Social Support | 273 | 0.7 (0.5, 1.0) | 0.06 |

HR: Hazards ratio. CI: confidence interval.
All models adjusted for age, sex, race (black versus non-black), married vs. not, education >12 years, ever smoking, BMI, history of hypertension, history of diabetes mellitus, left ventricular ejection fraction, and ST-segment elevation MI.
\*Daily mean ratings averaged across the week.

Next, we carried out a relative importance analysis by means of likelihood ratio chi-square tests (**Table 4**). In the model adjusted for demographic factors, medical history and clinical variables, the MNA ranked second of all variables in the model, after left ventricular ejection fraction, with a partial likelihood ratio χ² of 10.3 (p=0.001). In contrast, the global baseline negative affect score had a partial likelihood ratio of 3.4 (p=0.06). When both were included together in the same model, the partial likelihood ratio χ² for the MNA was only slightly reduced to 6.8 (p=0.009), while the partial likelihood ratio χ² for the baseline affect score went down to 0 (**Table 4**).

**Table 4.** Relative importance of momentary assessments of negative affect in daily life vs. negative affect measured at baseline for the prediction of major adverse cardiovascular events (cardiovascular death, nonfatal MI, stroke or hospitalizations for heart failure).

|  | HR (95% CI) Per SD Increment | P Value | Partial Likelihood Ratio Test† |  |
| --- | --- | --- | --- | --- |
| | | | $\chi^2$ (1 DF) | P Value |
| **Separately Added to Models with Covariates\*** |  |  |  |  |
| Momentary Negative Affect Score | 1.6 (1.2, 2.0) | 0.0004 | 10.3 | 0.001 |
| Baseline Negative Affect Score | 1.3 (0.9, 1.9) | 0.10 | 3.4 | 0.06 |
| **Jointly Added to Same Model with Covariates\*** |  |  |  |  |
| Momentary Negative Affect Score | 1.6 (1.2, 2.1) | 0.002 | 6.8 | 0.009 |
| Baseline Negative Affect Score | 1.0 (0.7, 1.4) | 0.97 | 0.0 | 0.97 |

HR: Hazards ratio. CI: confidence interval.
\*Age, sex, race (black versus non-black), married vs. not, education >12 years, ever smoking, BMI, history of hypertension, history of diabetes mellitus, left ventricular ejection fraction, and ST-segment elevation MI.
† The partial likelihood ratio chi-square statistic was calculated from the multivariable Cox proportional hazards model. It compares the $-2 \log$ partial likelihood of the full model to that of a reduced model omitting only that covariate. The resulting difference, distributed as a chi-square statistic with 1 degree of freedom, reflects the variation in the likelihood of the outcome uniquely attributable to that covariate after adjustment for the remaining covariates in the model.

In sensitivity analyses we utilized baseline assessments of depression, including the Beck Depression Inventory score and lifetime history of major depression, in place of the global baseline negative affect score. When these measures were added to the model adjusted for demographic factors, medical history and clinical variables, the MNA remained significant (HR per SD increase, 1.4, 95% CI, 1.0, 1.8, p=0.02) with a partial likelihood ratio χ² of 4.0 (p=0.046), while neither baseline depression measures reached statistical significance (Beck Depression Inventory, HR, 1.4 per SD increase, 95% CI, 0.98, 1.9, p=0.06); lifetime history of major depression, HR, 1.8, 95% CI, 0.8, 4.2, p=0.14). As a whole, these results suggest a larger contribution of MNA to MACE risk compared to aggregated baseline psychological assessments, with prognostic information that goes above and beyond depression status at baseline.

## Discussion

In this study of individuals recently hospitalized for an MI, we used EMA to sample experiences prospectively in daily life and found that moment-to-moment negative affect, assessed multiple times a day over one week, was associated with an increased risk of CVD events. Each SD increment in MNA, the daily mean of negative affect, was associated with 60% higher risk of MACE. When the MNA score was dichotomized at the median, a MNA score above the median was associated with approximately threefold elevated MACE risk. This association was similar in women and men and was robust to adjustment for potential confounders. Notably, the association was stronger than that observed for a composite measure of psychological scales administered retrospectively at the baseline visit, and was also independent of baseline measures of depression, including a lifetime history of major depressive disorder and baseline depressive symptom level. These findings suggest that momentary negative mood, captured as it occurs in daily life, may reveal cardiovascular risk that retrospective assessments miss, with potential implications for secondary prevention.

To our knowledge, ours is the first study of this kind in individuals with MI, but our findings are consistent with prior work in community samples indicating that real-time assessments at multiple time points can outperform retrospective measures in predicting cardiovascular health. For example, EMA ratings of high demand and low control were related to subclinical atherosclerosis assessed by intima-media thickness using carotid ultrasonography cross-sectionally and longitudinally, while traditional questionnaires of job strain did not show an association.^27,28^ Other studies have found only poor to modest agreement between real-time EMA tracking of mood and retrospective recall.^29^ In our study, EMA showed a stronger independent association with outcome than a composite measure of traditional retrospective scales measuring similar domains, even though the two types of measures were moderately correlated.

A potential reason for the stronger association of momentary assessments in daily life with CVD outcomes, compared with baseline psychological scales, is that gathering data as people go about their daily lives offers a more accurate assessment of subjective experiences than retrospective methodologies.^9^ Assessment methods that require participants to recall psychological states over the previous weeks or months have been criticized for potential recall bias and memory distortions.^7,13^ Furthermore, such retrospective assessments are disconnected from the natural context known to shape subjective experience. By contrast, EMA collects data prospectively, in real time, within participants’ natural environments through frequent repeated sampling, which enhances ecological validity and reduces the potential for recall bias.^7,13,30^ Another advantage of EMA is that averaging multiple measures over the week reduces random error and therefore produces more stable assessments that may help with prognostic significance.

Momentary positive affect showed a small protective relationship, but the estimate was not statistically significant. This pattern is consistent with previous prospective studies in predominantly healthy populations, in which negative mood has been more consistently linked to CVD risk than positive mood, including recalled mood^31^ and diurnal mood patterns.^13^ Momentary perceived stress also was not associated with MACE in our data, possibly because the assessment of stress was based on a single question of whether anything had happened in the previous hour that most people would consider stressful. It is likely that this measure of moment-to-moment stress is susceptible to variability in subjective interpretation. Momentary reports of stress were correlated with momentary mood, suggesting that at least in part, negative mood captured affective responses to stressful experiences.

There are multiple potential mechanisms underlying the association between daily negative affect and MACE. In previous research, depression and other negative psychological states have been associated with unhealthy lifestyle behaviors such as smoking, unhealthy diet and physical inactivity.^32^ Mood may also influence autonomic nervous system and hypothalamic-pituitary-adrenal axis functioning,^4^ and cardiac autonomic measures are known predictors of total and CVD mortality in the general population and in samples of people with CVD.^33,34^ Inflammation, platelet activation, and abnormal vascular function have also been implicated; ^35–37^ these are all mechanisms involved in the development of atherosclerosis or in the triggering of CVD events.^8^ In individuals with established CVD, mood states, of which depression has been the most studied, can also trigger silent ischemia^38,39^ and cardiac arrhythmias.^40^ It has also been argued that negative affect could be the expression of disease severity, which may explain why depression predicts worse outcomes in people with CVD.^4^ In our study, however, clinical severity characteristics were similarly distributed in participants below or above the median MNA score, and adjustment for clinical risk indicators did not affect the study estimate, suggesting that severity of disease was not a confounding factor in our analysis. Also, the relationship of MNA with MACE was independent of depression status at baseline, suggesting that EMA of negative affect captures risk beyond baseline depression.

As all the previous studies discussed above were based on retrospective measures of mood states, there is little information on mechanisms of risk for mood states specifically measured by EMA. Studies using wearable devices have generally confirmed that everyday stressors, psychological arousal, tension and anxiety are related to measures of cardiovascular reactivity.^41^ Most studies have used ambulatory blood pressure monitoring^42,43^ and ambulatory electrocardiographic monitoring.^44–46^ However, the recording time was often short and sample sizes were small. Most investigations also included healthy volunteers, and no studies examined high-risk populations such as those with a previous MI. Recent advances in mobile health technology may soon allow the unobtrusive collection and integration of a large amount of physiological data in conjunction with daily experiences, including passive ambulatory sensing of hemodynamic, vascular, autonomic and immune physiology. Future prospective studies with a large sample size using these technologies may be able to clarify the potential mechanisms underlying the association between daily mood states and CVD events in post-MI patients.

This study may help address existing gaps in the management of post-MI patients. Although a growing body of evidence supports addressing psychosocial factors in patients with CVD, the majority of class 1 secondary prevention guidance focuses on traditional risk factors and antiplatelet medications; screening and management of mental health is currently class 2a recommendation.^47^ Our data underscore the importance of capturing the psychological domain and suggest that we should transition beyond clinic settings to capture real-life situations. Wearable devices could also become invaluable in assessing responses to interventions. The standardization, validity and reproducibility of these techniques, however, need further evaluation.

This study has several limitations. Selection bias cannot be excluded, since patients who were too ill or otherwise unable to participate may have been underrepresented; this is inevitable in studies of post-MI patients. Missing responses to EMA prompts is another potential source of selection bias, although EMA missingness was only weakly correlated with daily mood. Because the correlation was negative, it is likely that the missingness led to underestimation of the true effect size. EMA also cannot be considered an “objective” measure, since it requires some degree of cognitive appraisal. However, subjective psychological responses may be as important to cardiovascular risk as objective characteristics.^48^ Furthermore, EMA administered in real time and in the natural environment should be less influenced by heuristic biases and memory errors than retrospective self-reports.^7^ The relatively small sample size is another limitation, and power for some analyses may have been affected. As our study was conducted at a single institution, the generalizability of our findings to other populations and settings requires further exploration. Despite these limitations, this study is innovative as we are not aware of other studies that have examined the link between moment-to-moment negative affect in daily life and risk of subsequent CVD events among individuals with a recent MI.

In conclusion, among young and midlife survivors of an MI, a greater burden of negative mood captured during everyday living was robustly and independently related to adverse CVD outcomes and was a stronger predictor than a battery of retrospective psychological measures obtained at the baseline visit. These findings underscore the contribution of emotional factors to morbidity and mortality after MI, with potentially important implications for secondary prevention. Tracking emotional experiences as they unfold in daily life may uncover cardiovascular risk that traditional risk indicators, and even baseline depression status, fail to capture. This technique could also help with monitoring the effects of behavioral interventions. Further studies are needed to establish the feasibility and utility of the EMA approach in routine clinical care. While treatment of depression after MI has not been shown to alter MI recurrence and mortality,^49,50^ clinical trials should test whether ameliorating daily mood states might help improve post-MI outcomes.

## Data Availability

The data that support the findings of this study are available from the corresponding author upon reasonable request.

## Supplemental Material

Supplemental Tables 1-3, Supplemental Figure 1.

## Non-standard Abbreviations and Acronyms

CAD: Coronary artery disease
CVD: Cardiovascular disease
DSM: Diagnostic Statistical Manual of Mental Disorders
EMA: Ecological momentary assessment
MI: Myocardial infarction
MIMS-3: Myocardial Infarction and Mental Stress Study 3
SCID: Structured Clinical Interview

## References

1. Garcia M, Moazzami K, Almuwaqqat Z, Young A, Okoh A, Shah AJ, Sullivan S, Lewis TT, Elon L, Ko YA, et al. Psychological Distress and the Risk of Adverse Cardiovascular Outcomes in Patients With Coronary Heart Disease. JACC Adv. 2024;3:100794. doi: 10.1016/j.jacadv.2023.100794

2. Gaffey AE, Gathright EC, Fletcher LM, Goldstein CM. Screening for Psychological Distress and Risk of Cardiovascular Disease and Related Mortality: A Systematized Review, Meta-analysis, and Case for Prevention. Journal of cardiopulmonary rehabilitation and prevention. 2022;42:404–415. doi: 10.1097/HCR.0000000000000751

3. Zeng J, Qiu Y, Yang C, Fan X, Zhou X, Zhang C, Zhu S, Long Y, Hashimoto K, Chang L, et al. Cardiovascular diseases and depression: A meta-analysis and Mendelian randomization analysis. Mol Psychiatry. 2025;30:4234–4246. doi: 10.1038/s41380-025-03003-2

4. Carney RM, Freedland KE. Depression and coronary heart disease. Nat Rev Cardiol. 2017;14:145–155. doi: 10.1038/nrcardio.2016.181

5. Kronish IM, Rieckmann N, Burg MM, Alcantara C, Davidson KW. The psychosocial context impacts medication adherence after acute coronary syndrome. nn Behav Med. 2014;47:158–164. doi: 10.1007/s12160-013-9544-0

6. Jones MC, Smith K, Herber O, White M, Steele F, Johnston DW. Intention, beliefs and mood assessed using electronic diaries predicts attendance at cardiac rehabilitation: An observational study. Int J Nurs Stud. 2018;88:143–152. doi: 10.1016/j.ijnurstu.2018.08.015

7. Kamarck TW, Schwartz JE, Shiffman S, Muldoon MF, Sutton-Tyrrell K, Janicki DL. Psychosocial stress and cardiovascular risk: what is the role of daily experience? J Pers. 2005;73:1749–1774. doi: 10.1111/j.0022-3506.2005.00365.x

8. Vaccarino V, Shah AJ, Mehta PK, Pearce B, Raggi P, Bremner JD, Quyyumi A. Stress response physiology in the cardiac patient: from the laboratory to real life. Stress. 2026;29:2723352. doi: 10.1080/10253890.2026.2723352

9. Smyth JM, Zawadzki MJ, Marcusson-Clavertz D, Scott SB, Johnson JA, Kim J, Toledo MJ, Stawski RS, Sliwinski MJ, Almeida DM. Computing Components of Everyday Stress Responses: Exploring Conceptual Challenges and New Opportunities. Perspect Psychol Sci. 2023;18:110–124. doi: 10.1177/17456916221082108

10. Wrzus C, Neubauer AB. Ecological Momentary Assessment: A Meta-Analysis on Designs, Samples, and Compliance Across Research Fields. Assessment. 2023;30:825–846. doi: 10.1177/10731911211067538

11. Jean FAM, Sibon I, Husky M, Couffinhal T, Swendsen J. Feasibility and validity of Ecological Momentary Assessment in patients with acute coronary syndrome. BMC Cardiovasc Disord. 2020;20:499. doi: 10.1186/s12872-020-01774-w

12. Vaccarino V, She H, Elon L, Lewis TT, Ko YA, Clifford G, Li Q, Murrah N, Shallenberger LH, Roberts T, et al. Real-life Psychological Experiences and Cardiac Autonomic Physiology among Survivors of a Myocardial Infarction in Midlife. JAHA - in press. Pre print available at. Res Sq. 2025. doi: 10.21203/rs.3.rs-7418040/v1

13. Kroenke CH, Seeman T, Matthews K, Adler N, Epel E. Mood patterns based on momentary assessment of positive and negative moods over a day and coronary artery calcification in the CARDIA study. Psychosom Med. 2012;74:526–534. doi: 10.1097/PSY.0b013e3182583e68

14. Thygesen K, Alpert JS, Jaffe AS, Chaitman BR, Bax JJ, Morrow DA, White HD, Executive Group on behalf of the Joint European Society of Cardiology /American College of Cardiology /American Heart Association /World Heart Federation Task Force for the Universal Definition of Myocardial I. Fourth Universal Definition of Myocardial Infarction (2018). Circulation. 2018;138:e618–e651. doi: 10.1161/CIR.0000000000000617

15. First MB, Spitzer RL, Williams JBW, Gibbon M. Structured Clinical Interview for DSM IV-Patient Edition (SCID-P). Washington, D.C.: American Psychiatric Press; 1995.

16. Beck AT, Steer RA, Brown GK. BDI-II. Beck Depression Inventory: Second Edition. San Antonio, TX: The Psychological Corporation; 1996.

17. Spielberger CD, Gorsuch RL, Lushene RE. State-Trait Anxiety (STAI) manual. Palo Alto, CA: Consulting Psychologists Press; 1970.

18. Spielberger CD. Professional manual for the State-Trait Anger Expression Inventory. Research Ed., Tampa, Fla: University of South Florida; 1988.

19. Cohen S, Kamarck T, Mermelstein R. A global measure of perceived stress. J Health Soc Behav. 1983;24:385–396.

20. Canty-Mitchell J, Zimet GD. Psychometric properties of the Multidimensional Scale of Perceived Social Support in urban adolescents. Am J Community Psychol. 2000;28:391–400. doi: 10.1023/A:1005109522457

21. Neeland IJ, Patel RS, Eshtehardi P, Dhawan S, McDaniel MC, Rab ST, Vaccarino V, Zafari AM, Samady H, Quyyumi AA. Coronary angiographic scoring systems: an evaluation of their equivalence and validity. Am Heart J. 2012;164:547–552 e541. doi: 10.1016/j.ahj.2012.07.007

22. Stawski RS, Cichy KE, Piazza JR, Almeida DM. Associations among daily stressors and salivary cortisol: findings from the National Study of Daily Experiences. Psychoneuroendocrinology. 2013;38:2654–2665. doi: 10.1016/j.psyneuen.2013.06.023

23. Sin NL, Graham-Engeland JE, Ong AD, Almeida DM. Affective reactivity to daily stressors is associated with elevated inflammation. Health Psychol. 2015;34:1154–1165. doi: 10.1037/hea0000240

24. Amrhein V, Greenland S, McShane B. Retire statistical significance. Nature. 2019;567:305–307.

25. Rosenbaum PR, Rubin DB. The central role of the propensity score in observational studies for causal effects. Biometrika. 1983;70:41–55. doi: 10.1093/biomet/70.1.41

26. Hirano K, Imbens GW. The Propensity Score with Continuous Treatments. In: Applied Bayesian Modeling and Causal Inference from Incomplete-Data Perspectives. 2004:73–84.

27. Kamarck TW, Muldoon MF, Shiffman SS, Sutton-Tyrrell K. Experiences of demand and control during daily life are predictors of carotid atherosclerotic progression among healthy men. Health Psychol. 2007;26:324–332. doi: 10.1037/0278-6133.26.3.324

28. Kamarck TW, Muldoon MF, Shiffman S, Sutton-Tyrrell K, Gwaltney C, Janicki DL. Experiences of demand and control in daily life as correlates of subclinical carotid atherosclerosis in a healthy older sample. Health Psychol. 2004;23:24–32. doi: 10.1037/0278-6133.23.1.24

29. Solhan MB, Trull TJ, Jahng S, Wood PK. Clinical assessment of affective instability: comparing EMA indices, questionnaire reports, and retrospective recall. Psychological assessment. 2009;21:425–436. doi: 10.1037/a0016869

30. Doherty K, Balaskas A, Doherty G. The Design of Ecological Momentary Assessment Technologies. Interacting with Computers. 2020;32:257–278. doi: 10.1093/iwcomp/iwaa019

31. Nabi H, Kivimaki M, De Vogli R, Marmot MG, Singh-Manoux A. Positive and negative affect and risk of coronary heart disease: Whitehall II prospective cohort study. BMJ. 2008;337:a118. doi: 10.1136/bmj.a118

32. Johnstad PG. Unhealthy behaviors associated with mental health disorders: a systematic comparative review of diet quality, sedentary behavior, and cannabis and tobacco use. Frontiers in Public Health. 2024;Volume 11 - 2023. doi: 10.3389/fpubh.2023.1268339

33. Bauer A, Kantelhardt JW, Barthel P, Schneider R, Makikallio T, Ulm K, Hnatkova K, Schomig A, Huikuri H, Bunde A, et al. Deceleration capacity of heart rate as a predictor of mortality after myocardial infarction: cohort study. Lancet. 2006;367:1674–1681. doi: 10.1016/S0140-6736(06)68735-7

34. Jarczok MN, Weimer K, Braun C, Williams DP, Thayer JF, Gundel HO, Balint EM. Heart rate variability in the prediction of mortality: A systematic review and meta-analysis of healthy and patient populations. Neurosci Biobehav Rev. 2022;143:104907. doi: 10.1016/j.neubiorev.2022.104907

35. Osimo EF, Pillinger T, Rodriguez IM, Khandaker GM, Pariante CM, Howes OD. Inflammatory markers in depression: A meta-analysis of mean differences and variability in 5,166 patients and 5,083 controls. Brain Behav Immun. 2020;87:901–909. doi: 10.1016/j.bbi.2020.02.010

36. Zafar MU, Paz-Yepes M, Shimbo D, Vilahur G, Burg MM, Chaplin W, Fuster V, Davidson KW, Badimon JJ. Anxiety is a better predictor of platelet reactivity in coronary artery disease patients than depression. Eur Heart J. 2010;31:1573–1582. doi: 10.1093/eurheartj/ehp602

37. Shimbo D, Child J, Davidson K, Geer E, Osende JI, Reddy S, Dronge A, Fuster V, Badimon JJ. Exaggerated serotonin-mediated platelet reactivity as a possible link in depression and acute coronary syndromes. Am J Cardiol. 2002;89:331–333. doi: 10.1016/s0002-9149(01)02236-6

38. Wei J, Pimple P, Shah AJ, Rooks C, Bremner JD, Nye JA, Ibeanu I, Murrah N, Shallenberger L, Raggi P, et al. Depressive symptoms are associated with mental stress-induced myocardial ischemia after acute myocardial infarction. PloS one. 2014;9:e102986. doi: 10.1371/journal.pone.0102986

39. Boyle SH, Samad Z, Becker RC, Williams R, Kuhn C, Ortel TL, Kuchibhatla M, Prybol K, Rogers J, O’Connor C, et al. Depressive symptoms and mental stress-induced myocardial ischemia in patients with coronary heart disease. Psychosom Med. 2013;75:822–831. doi: 10.1097/PSY.0b013e3182a893ae

40. Lampert R, Joska T, Burg MM, Batsford WP, McPherson CA, Jain D. Emotional and physical precipitants of ventricular arrhythmia. Circulation. 2002;106:1800–1805. doi: 10.1161/01.cir.0000031733.51374.c1

41. Whittaker AC, Ginty A, Hughes BM, Steptoe A, Lovallo WR. Cardiovascular Stress Reactivity and Health: Recent Questions and Future Directions. Psychosom Med. 2021;83:756–766. doi: 10.1097/PSY.0000000000000973

42. Kamarck TW, Janicki DL, Shiffman S, Polk DE, Muldoon MF, Liebenauer LL, Schwartz JE. Psychosocial demands and ambulatory blood pressure: a field assessment approach. Physiol Behav. 2002;77:699–704. doi: 10.1016/s0031-9384(02)00921-6

43. Raikkonen K, Matthews KA. Do dispositional pessimism and optimism predict ambulatory blood pressure during school days and nights in adolescents? J Pers. 2008;76:605–630. doi: 10.1111/j.1467-6494.2008.00498.x

44. Szakonyi B, Vassanyi I, Schumacher E, Kosa I. Efficient methods for acute stress detection using heart rate variability data from Ambient Assisted Living sensors. Biomed Eng Online. 2021;20:73. doi: 10.1186/s12938-021-00911-6

45. Rodrigues S, Paiva JS, Dias D, Cunha JPS. Stress among on-duty firefighters: an ambulatory assessment study. PeerJ. 2018;6:e5967. doi: 10.7717/peerj.5967

46. Smets E, Rios Velazquez E, Schiavone G, Chakroun I, D’Hondt E, De Raedt W, Cornelis J, Janssens O, Van Hoecke S, Claes S, et al. Large-scale wearable data reveal digital phenotypes for daily-life stress detection. NPJ Digit Med. 2018;1:67. doi: 10.1038/s41746-018-0074-9

47. Writing Committee M, Virani SS, Newby LK, Arnold SV, Bittner V, Brewer LC, Demeter SH, Dixon DL, Fearon WF, Hess B, et al. 2023 AHA/ACC/ACCP/ASPC/NLA/PCNA Guideline for the Management of Patients With Chronic Coronary Disease: A Report of the American Heart Association/American College of Cardiology Joint Committee on Clinical Practice Guidelines. J Am Coll Cardiol. 2023;82:833–955. doi: 10.1016/j.jacc.2023.04.003

48. Vaccarino V, Bremner JD. Stress and cardiovascular disease: an update. Nature reviews Cardiology. 2024;21:603–616. doi: 10.1038/s41569-024-01024-y

49. Berkman LF, Blumenthal J, Burg M, Carney RM, Catellier D, Cowan MJ, Czajkowski SM, DeBusk R, Hosking J, Jaffe A, et al. Effects of treating depression and low perceived social support on clinical events after myocardial infarction: the Enhancing Recovery in Coronary Heart Disease Patients (ENRICHD) Randomized Trial. JAMA. 2003;289:3106–3116. doi: 10.1001/jama.289.23.3106

50. van Melle JP, de Jonge P, Honig A, Schene AH, Kuyper AMG, Crijns HJGM, Schins A, Tulner D, van den Berg MP, Ormel J. Effects of antidepressant treatment following myocardial infarction. The British Journal of Psychiatry. 2007;190:460–466. doi: 10.1192/bjp.bp.106.028647

